# The APOL1 protective M1 modifier occurs exclusively with the KIK protein haplotype to suppress channel conductance

**DOI:** 10.64898/2026.09.14.26362999

**Authors:** Diya Yang, Korey R. Bartolomeo, Shiying Liu, Agustin Gonzalez-Vicente, Nicholas R. Wheeler, Penelope Benchek, Xiaofeng Zhu, Jonathan L. Haines, Jessica N. Cooke Bailey, Thomas Nader, Nurdan Cam, Leslie A. Bruggeman, Scott M. Williams, Dana C. Crawford, William S. Bush, John F. O’Toole, John R. Sedor

**Affiliations:** Department of Population and Quantitative Health Sciences, School of Medicine, Case Western Reserve University, Cleveland, OH, USA; Cleveland Institute for Computational Biology, Case Western Reserve University, Cleveland, OH, USA; Department of Genetics and Genome Sciences, School of Medicine, Case Western Reserve University, Cleveland, OH, USA; Department of Physiology and Biophysics, School of Medicine, Case Western Reserve University, Cleveland, OH, USA; Department of Molecular Medicine, Cleveland Clinic Lerner College of Medicine of Case Western Reserve University School of Medicine, Cleveland, OH, USA; Department of Pharmacology & Toxicology, East Carolina University, Greenville, NC, USA; Department of Kidney Medicine, Medical Specialties Institute and Department of Heart, Blood and Kidney Research, Cleveland Clinic, Cleveland, OH, USA

**Keywords:** *APOL1*, p.N264K, haplotype, cation channel, genetic ancestry, chronic kidney disease

## Abstract

**Introduction:** Genetic studies have shown the *APOL1* variant p.N264K, also called M1, is associated with markedly lower kidney disease risk when co-inherited with the G2-risk allele. Empirical studies show M1 in *cis* with G2 blocks APOL1 cation channel flux. To date M1 only occurs on a specific APOL1 haplotype defined by three amino acid positions: p.Lys150 (K), p.Ile228 (I) and p.Lys255 (K). We analyzed two large community datasets to determine if M1 occurs on other APOL1 protein haplotypes and used a cell model to assess if the M1 variant alone is sufficient to block channel activity or if its haplotype context modifies its function.

**Methods:** Phased *APOL1* haplotypes frequencies were examined in the Alzheimer Disease Sequencing Project Release 5 (ASDP R5) participants (n=40,909) and confirmed in All of Us Research Program (All of Us) enrollees (n=72,538). Reported race was compared with genetically inferred ancestry. Local APOL1 genealogies were inferred and allele age determined. Thallium flux was measured in 293 cells after induced expression of *APOL1*-G2 on natural and engineered haplotype backgrounds.

**Results:** The M1 variant occurred exclusively on the KIK APOL1 protein haplotype in all ADSP R5 participants (n=330) and 99.5% of All of Us participants (n=3,560). Local genealogies placed these chromosomes within the lineage carrying p.150K. *In vitro* thallium flux assays demonstrated that M1 alone and p.Lys150 alone each partially reduced thallium flux but completely abrogated thallium flux when on the same haplotype.

**Conclusions:** M1 is inherited almost exclusively with a single haplotype background. Both the p.150K and p.264K (M1) are needed to completely block APOL1-G2-mediated thallium flux. Studies aimed at examining and exploiting the protective mechanisms of M1 for possible treatments need to consider its distinct haplotype context.

**Translational Statement:** G1 and G2 coding variants in *APOL1* increase the risk of kidney disease in populations with African ancestry. Another coding variant p.N264K (M1) reduces kidney disease risk when inherited in *cis* with the APOL1 G2 risk allele. Here we show in large populations that the M1 is almost always inherited on an *APOL1* haplotype that encodes a p.E150K amino acid change. Otherwise G2 in the absence of M1 is exclusively on a p.150E haplotype. M1 and p.150K functionally interact to block APOL1-mediated thallium flux in cell culture models. These functional interactions highlight the importance of considering the M1 variant in its haplotype context when designing basic research studies and translating results into future approaches for therapeutic intervention.

## Introduction

Much of the excess biological risk for kidney disease in Black people is explained by the association of two nonsynonymous coding variants, G1 and G2, in the *APOL1* gene, which are found exclusively in people of African ancestry.^1,2^ *APOL1* has ion channel activity and both G1 and G2 prevent African sleeping sickness caused by the *Trypanosoma brucei* subspecies that infect humans. The *APOL1* locus has additional coding polymorphisms that define distinct common haplotypes.^3–6^ G0, *APOL1* without kidney risk variants, is found on a number of common haplotypes defined, in part, by variation at p.E150K, p.M228I and p.R255K (KIK) (**Figure S1, Table S1**).^3,4^ In contrast, the kidney disease risk variants G1 and G2 segregate almost exclusively with p150E, p228I and p255K. In *vitro* studies demonstrate that the haplotype context modifies *APOL1* cytotoxicity and ion channel conductance.^6–8^

Prior studies have demonstrated that a low-frequency *APOL1* coding variant (p.N264K [M1]), when inherited with the G2 haplotype, reduces risk for chronic kidney disease, kidney failure and focal segmental glomerulosclerosis,^9–11^ an effect attributed to occlusion of the G2 APOL1 channel with loss of cation flux and resulting blunted cytotoxicity.^9,12,13^ M1 is never inherited with G1 in *cis*. A haplotype analysis of 1000 Genomes data found M1 only on a KIK haplotype, with either G0 or G2, and proposed that M1-G2 arose by recombination between an M1-bearing KIK chromosome and an EIK G2 chromosome.^14^ Our study has two goals (**Figure S1**): first, to examine *APOL1* haplotype frequencies including p264 in two large population datasets not ascertained for kidney disease; and second, to assess whether the haplotype context of M1 modifies its effect on *APOL1* G2 ion channel activity in a cell culture model system.

## Short Methods

*APOL1* haplotypes are defined by a four-letter code giving the amino acids at positions 150, 228, 255 and 264, the first three being background positions previously reported^3–6^ and the fourth distinguishing ancestral asparagine 264 from the derived lysine encoded by the M1 modifier p.N264K (rs73885316), followed by a suffix indicating the absence (G0) or presence (G1 or G2) of a kidney risk allele *in cis* (as shown in **Table S1, Figure S1**). APOL1 haplotype frequencies were determined in participants of the Alzheimer Disease Sequencing Project Release 5 (ADSP R5, n=40,909 participants)^15^ and confirmed in participants of the All of Us Research Program (All of Us, n=72,538).^16^ Neither population was ascertained for kidney disease so the haplotypes frequencies reflect community prevalence. *APOL1* channel conductance was measured with the FluxOR II Green Potassium Ion Channel Assay (Thermo Fisher) in HEK 293 cells stably expressing tetracycline-regulatable *APOL1* G2 transgenes with EIKN, EIKK, KIKN and KIKK haplotypes. Complete methods are provided in the **Supplementary Materials**.

## Results

We characterized phased *APOL1* haplotypes in 40,909 ADSP R5 participants (81,818 chromosomes). The 4,371 participants reported as Black or African American (8,742 chromosomes) in ADSP were the initial population used to determine distribution of *APOL1* haplotype variation (**Table 1**). Background haplotype frequencies in the Black or African American ADSP R5 dataset are consistent with *APOL1* haplotype distributions reported previously for African-ancestry samples^8^ and different from the frequencies for other ADSP R5 reported-race categories in **Table 2**. G1 was present on 22.13% and G2 on 12.42% of chromosomes and the M1 modifier p.N264K was on 2.10% of the sample. G1 and G2 were confined to the EIK background throughout.

**Table 1.** *APOL1* Haplotype Frequencies in Participants Reported as Black or African American in ADSP R5 and All of Us.

| Haplotype | ADSP R5 (N = 4,371;<br>chromosomes = 8,742)<br>n (%) | AoU (N = 72,538;<br>chromosomes = 145,076)<br>n (%) |
| --- | --- | --- |
| KIKN_G0 | 2,973 (34.01) | 47,988 (33.08) |
| EIKN_G0 | 2,111 (24.15) | 34,826 (24.01) |
| EIKN_G1 | 1,935 (22.13) | 31,948 (22.02) |
| EIKN_G2 | 1,025 (11.73) | 18,763 (12.93) |
| EMRN_G0 | 458 (5.24) | 6,678 (4.60) |
| KIKK_G0 | 125 (1.43) | 2,223 (1.53) |
| KIKK_G2 | 59 (0.67) | 1,337 (0.92) |
| EMKN_G0 | 52 (0.59) | 1,120 (0.77) |
| Other labeled haplotypes | 4 (0.05) | 193 (0.13) |

**Table 2.**
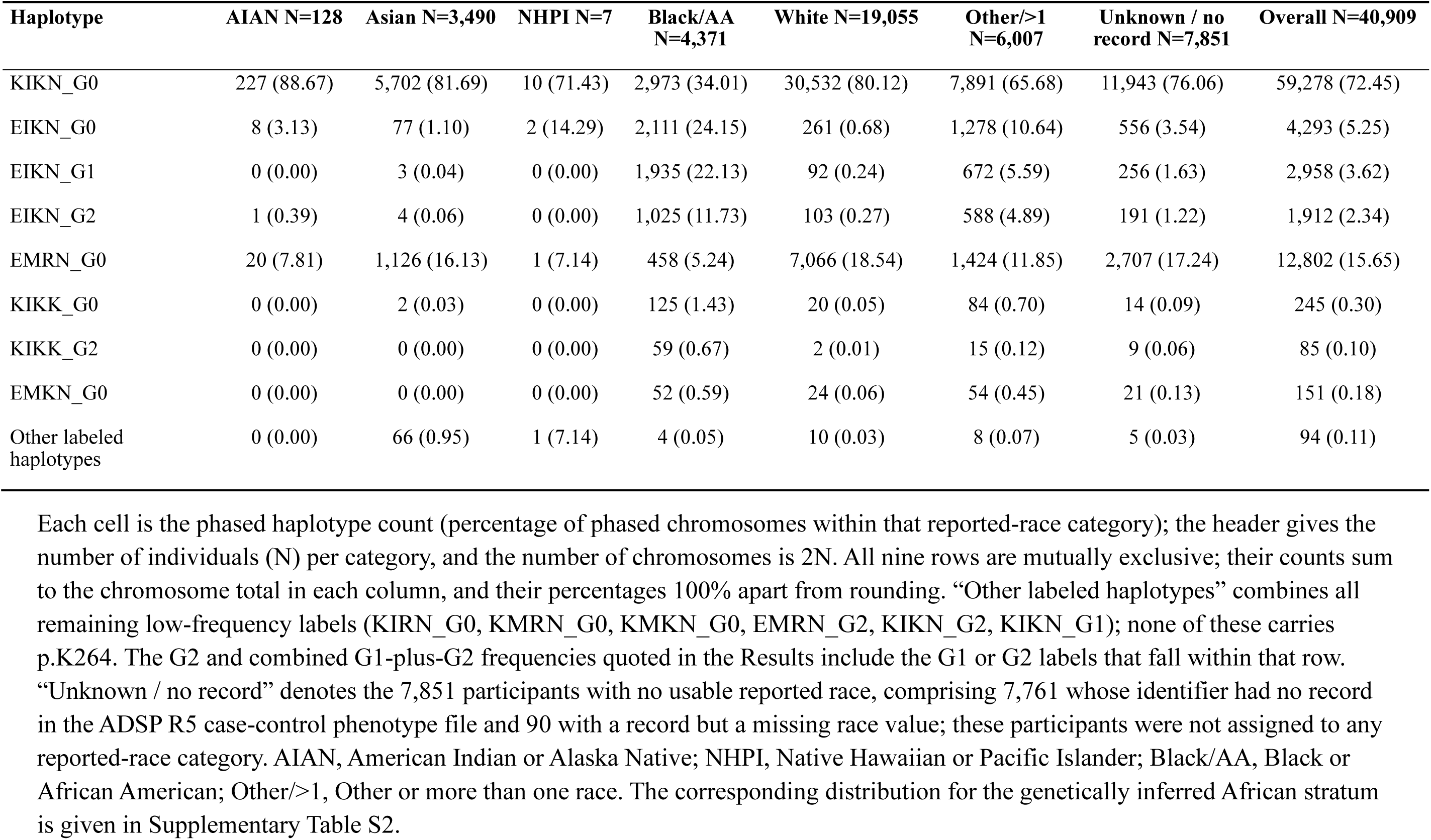
Distribution of *APOL1* Haplotypes by ADSP R5 Reported-Race Category.

Frequencies among the ADSP African descent individuals were very similar to those of 72,538 All of Us participants reporting non-Hispanic Black race (**Table 1**; r = 0.999, largest difference 1.21% points). KIKN_G0 was the most common single haplotype in both study populations (34.01% and 33.08%), and the background EIK APOL1 protein haplotype predominated (58.01% and 58.96%). In ADSP R5 every haplotype carrying M1 (lysine 264) had a KIK background. All 330 p.K264 chromosomes across reported-race strata were KIKK, with none on another background haplotype or *in cis* with G1 (**Tables 1** and **2**). In the African ancestry stratum, the prevalence of KIKK_G0 (n = 125) was 1.43% and KIKK_G2 (n = 59) was 0.67%, similar to prior reports.^9,10^ All of Us showed the same pattern: more than 99.5% of M1 chromosomes were KIK (KIKK_G0, 1.53%; KIKK_G2, 0.92%) (**Table 2**). Larger sample sizes are needed to confidently determine if the few non-KIK-M1 chromosomes identified were valid or technical errors.

The haplotype distributions in the ADSP R5 4,455 participants with African continental genetic ancestry closely matched those obtained using reported-race (r = 0.998), and all 197 p.K264 chromosomes had KIKK haplotypes (**Supplementary Table S2**). Reported race agreed closely with genetic ancestry (Cohen kappa = 0.90; 97.8% agreement), and model-based clustering showed the same continental structure (**Supplementary Tables S3 and S4; Supplementary Figure S1**). M1, G1 and G2 carriers clustered nearest the 1000 Genomes African superpopulation; G1 frequency rose with African ancestry, whereas G2 and M1 frequencies were approximately constant (**Supplementary Figure S2**).

Local genealogies placed p.M228I on the deepest focal branch, followed by p.E150K, and nested p.N264K within the p.E150K-containing KIK lineage as a recent descendant clade, supporting a model in which p.N264K arose after p.E150K on that background and is compatible with the proposed recombinational formation of KIKK_G2 (**Supplementary Figures S3 and S4**).^14^ Because the genealogies were inferred under a neutral coalescent model at a locus with evidence of selection, we interpret these results as relative ordering rather than absolute allele ages.

To test whether the M1 block of G2 conductance depends on haplotype context, we measured thallium (Tl⁺) flux, a surrogate for potassium movement through the APOL1 channel,^17^ in tetracycline-inducible T-REx-293 cells expressing G2 *APOL1* on the human EIKN and KIKK and engineered EIKK and KIKN backgrounds (**Figure 1a**). Induced *APOL1* abundance in the EIKK, KIKN and KIKK lines matched or exceeded that in EIKN, so their lower flux is not explained by lower total APOL1.^18^ Relative to EIKN_G2, which had the strongest induced flux, p.K264 alone (EIKK_G2; Tukey-adjusted *P* < 0.01) and p.E150K alone (KIKN_G2; *P* < 0.001) each incompletely reduced flux to a similar extent. In contrast, thallium conductance in 293 cells with KIKK_G2 transgenes was completely suppressed and indistinguishable from untreated cells (**Figure 1b and 1c**). Inaxaplin, an *APOL1* channel inhibitor,^19,20^ blocked induced thallium flux. Nigericin, an ionophore that exchanges potassium for hydrogen ions across biologic membranes, stimulated thallium flux in all four cell lines.

**Figure 1.**
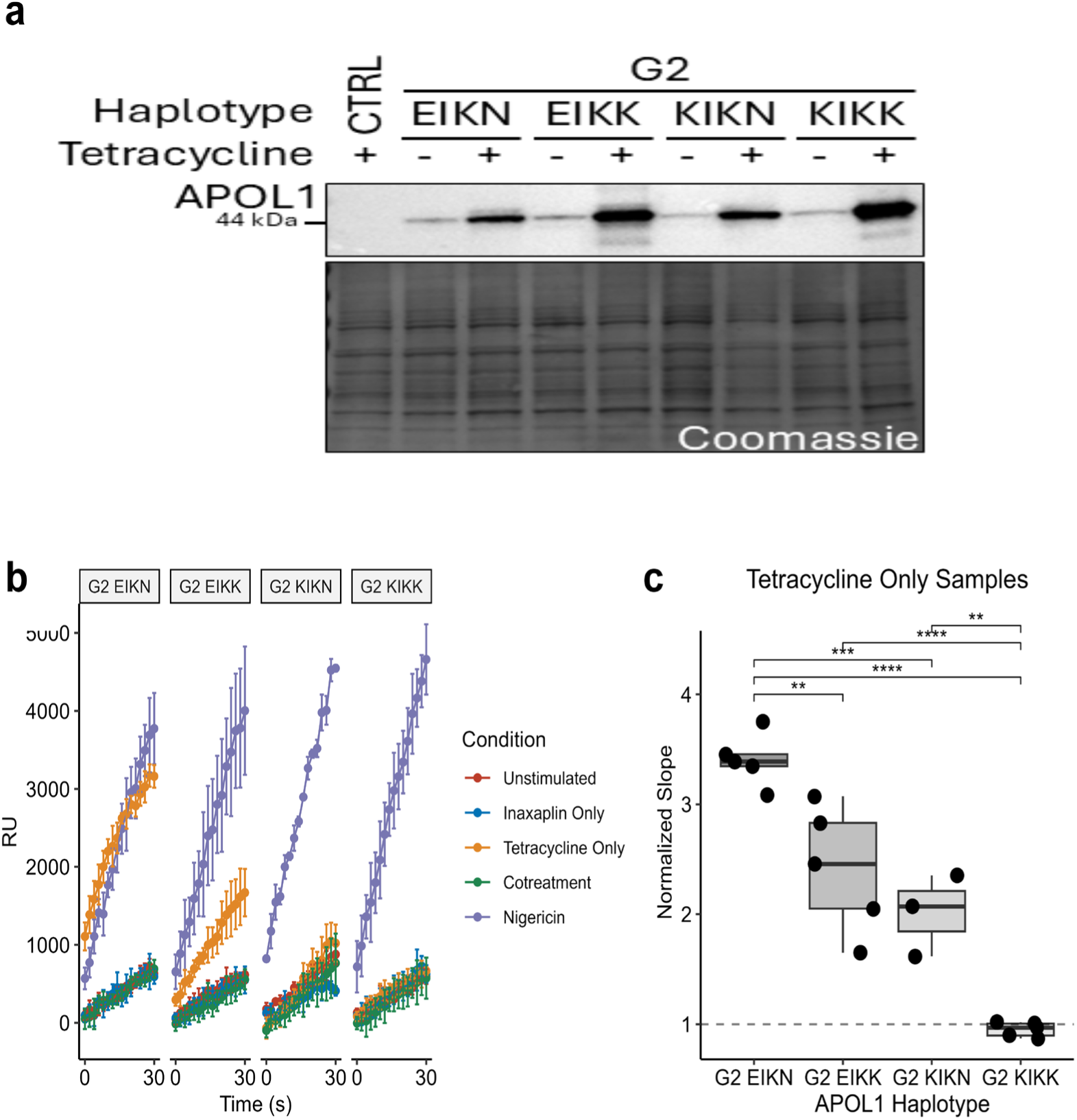
*APOL1* p.K150 and p.K264 combine to reduce G2-associated thallium flux. T-REx-293 cells with stable genomic integration of tetracycline-inducible G2 APOL1 transgenes on EIKN, EIKK, KIKN, or KIKK backgrounds were treated for 6 hours with tetracycline (100 ng/mL), as indicated. (A) Induced APOL1 abundance in EIKK, KIKN, and KIKK cells matched or exceeded that in EIKN cells. (B) Tl^+^ influx is shown as the change in relative fluorescence units after thallium addition. Relative to induced EIKN, flux was reduced but remained detectable with EIKK or KIKN and was reduced to the unstimulated baseline with KIKK. (C) Box plots show the slope in tetracycline-only cells divided by the slope in unstimulated cells of the same haplotype; the dashed line at 1 indicates no change from the uninduced baseline. n = 3-5 independent experiments. Two-way ANOVA with Tukey honestly significant difference test; **P < 0.01, ***P < 0.001, ****P < 0.0001; comparisons not shown were not significant. Inaxaplin, 3 µM; cotreatment, tetracycline plus inaxaplin; nigericin, 20 µM.

## Discussion

Across two large, independent United States datasets, p.N264K occurred almost exclusively on the *APOL1* KIK background: all 330 p.K264 chromosomes in ADSP R5 were KIKK, as were more than 99.5% in All of Us. Our Tl⁺ flux assays support a modulating effect of haplotype background on the proposed cation channel function of *APOL1*: p.K264 on EIK reduced but did not abolish activity, whereas KIKK reduced flux to the uninduced baseline. The genealogies place the background haplotype amino acid substitutions on deep branches and p.N264K as a recent clade inside the p.E150K lineage. A proximity-recombination origin has been proposed for KIKK_G2.^14^

Haplotype-aware reporting therefore matters for clinical risk models, where *APOL1* genotype is under prospective study,^21^ and for *APOL1*-directed therapy eligibility. Specifically, clinical testing should retain p.N264K because it identifies G2 chromosomes with much lower kidney disease risk.^10,11,22^ However, because it marks KIKK on essentially every chromosome carrying it, reporting it as an isolated causal modifier overstates what human data can resolve. Functional studies should report the complete background and distinguish natural KIKK from engineered EIKK constructs, since the cell-based studies of p.N264K have used an EIK background on which it is essentially absent in humans.^12^

This study characterized *APOL1* haplotype structure, not clinical outcomes as neither cohort had kidney phenotypes. Among Black or African American ADSP R5 participants G1, G2, M1 allele or kidney risk genotype frequencies were not statistically different between individuals with across no cognitive impairment, mild cognitive impairment and Alzheimer dementia, whereas *APOE* ε4 rose steeply (**Supplementary Table S5**). A separate multi-ancestry analysis found no significant genome-wide genetic correlation between kidney function and Alzheimer disease.^23^

We evaluated if APOL1 background protein haplotype modified the ability of the M1 to suppress APOL1 ion channel activity. M1 has previously been shown to block increases in intracellular calcium in cells with EIK-G2 transgenes.^9,13^ Since changes in cytosolic calcium have been shown to be downstream of APOL1 monovalent channel conductance,^24^ we measured Tl^+^ flux as a surrogate for potassium movement and found M1 required p150K to completely block channel activity. Interestingly, the rs2239785 coding variant for p.E150K has been nominally associated with reduced kidney disease risk.^5,25^ The basis for the p150K and P264K interaction is speculative given the absence of an empirical model for full length APOL1 structure. Both amino acid changes result in a gain in positive change near the first and third transmembrane domains, which may result in conformational changes that block channel activity. In conclusion we show in large populations not ascertained for kidney disease that the APOL1 protective M1 modifier occurs exclusively with the KIK background haplotype. The presence of p150K is required to completely block monovalent cation activity in cell models, highlighting the importance of haplotype context in future studies of M1 for future APOL1 kidney disease therapies.

## Supplementary Material

The supplementary material file includes Supplementary Methods, Supplementary Results, Supplementary Tables S1 to S5, Supplementary Figures S1 to S4, Supplementary Acknowledgements and Supplementary References and is available in supplement.

## Data Sharing Statement

ADSP data are available through the National Institute on Aging Genetics of Alzheimer’s Disease Data Storage Site (NIAGADS; https://www.niagads.org/) under the ADSP umbrella accession NG00067. Data were used under approved data use agreements with NIAGADS and dbGAP. All of Us data are available to registered investigators through the All of Us Researcher Workbench (https://www.researchallofus.org/). The All of Us data were accessed from the All of Us Research Program Curated Data Repository version 8 and are accessible through All of Us Controlled Tier access. Analysis code is available from the corresponding author on request.

## Ethics Statement

This study was approved by the Institutional Review Board at Case Western Reserve University. All included ADSP R5 participants provided written informed consent for General Research Use (GRU) or Health/Medical/Biomedical (HMB) purposes. All of Us Research Program participants provided informed electronic consent which included authorization for the broad secondary use of their de-identified data. The All of Us Research Program protocol is overseen by the All of Us Institutional Review Board which determined that research using the de-identified data through the Researcher Workbench is not human subjects research.

## Author Contributions

J.R.S. and J.F.O. had the original idea for the study. D.Y., J.R.S. and J.F.O. designed the work. D.Y. performed the genetic analyses on ADSP R5, curated and organized the data, produced the figures and tables, and drafted the manuscript. K.R.B. led the cell-based experimental work. K.R.B., T.N., N.C., L.A.B, J.O.T. and J.R.S. performed and/or interpreted the thallium flux and immunoblot experiments. S.L., J.C.C. and D.C.C. generated the All of Us haplotype frequency data. N.R.W. and P.B. generated and quality-controlled the ADSP R5 sequence data used here. A.G.-V. helped compile the variant annotation presented in Supplementary Table S1. X.Z., J.L.H. and S.M.W. advised on the genetic analyses and their interpretation. W.S.B. and D.C.C. contributed to development of the study concept and to interpretation of the genetic data. All authors critically revised the manuscript and approved the final version.

## Declaration of Generative AI and AI-Assisted Technologies in the Manuscript Preparation Process

Claude (Anthropic) and ChatGPT (OpenAI) were used solely for language editing (grammar, spelling, clarity, and conciseness). All scientific content, literature evaluation, analysis, interpretation of findings, and conclusions are entirely the authors’ original work. The authors reviewed and edited the content as needed and assume full responsibility for the published article.

## Acknowledgments

The authors sincerely thank the participants and investigators of the Alzheimer Disease Sequencing Project. We thank the National Institutes of Health’s All of Us Research Program for making available the participant data examined in this study. The *All of Us* Research Program would not be possible without the partnership of its participants to advance science and better health for all of us. Data for this study were prepared, archived, and distributed by the National Institute on Aging Alzheimer’s Disease Data Storage Site (NIAGADS) at the University of Pennsylvania (U24-AG041689), funded by the National Institute on Aging. Detailed acknowledgments for the Alzheimer’s Disease Sequencing Project are in supplementary materials.

