## Supplementary Files for "The APOL1 protective M1 modifier occurs exclusively with the KIK protein haplotype to suppress channel conductance"

### Supplementary Methods

#### 1. Genetic analyses

***Study datasets and participants***

***Alzheimer Disease Sequencing Project (ADSP).***

We analyzed ADSP Release 5 (ADSP R5). All seven ADSP R5 strata, that is the six reported-race categories plus participants with no usable reported race (40,909 participants; 81,818 phased chromosomes), were processed through the same pipeline, and haplotype frequencies for every stratum are reported in Table 2 of the main text. The primary analysis stratum was the 4,371 participants reported as Black or African American, and is the only reported-race category in which the G1 and G2 kidney risk alleles were common: 34.56% of chromosomes in that stratum carried G1 or G2, compared with 10.62% for Other or more than one race, 2.90% for participants with no usable reported race, 0.52% for White, 0.39% for American Indian or Alaska Native, 0.10% for Asian, and 0% for Native Hawaiian or Pacific Islander. The American Indian or Alaska Native stratum carried a single G2 chromosome and no G1 chromosome and is reported for completeness only. Participants were older adults ascertained for Alzheimer disease research, not for kidney disease, and the cohort is not population-based. Since ascertainment was blind to kidney risk variant status, the haplotype frequencies reported here are similar to community prevalence. Individuals, who limited the use of their data to dementia studies, were not included.

***All of Us Research Program***

Haplotype frequencies were computed from the All of Us Research Program (data release 8), whose genomic dataset is described elsewhere.[S1] Of more than 800,000 participants, approximately 414,000 had short-read whole-genome sequences, phased with Beagle.[S2] Frequencies reported here are for 72,538 participants who reported non-Hispanic Black race, corresponding to 145,076 phased chromosomes.

##### *Genotyping and phasing (ADSP)*

Genotypes were taken from ADSP R5 joint-genotyped whole-genome sequencing data for chromosome 22 on the GRCh38 build. Variant records were split and normalized, and multiallelic records were resolved. Each reported-race group was phased separately with Beagle 5.5[S2] using GRCh38 human genetic maps.

##### *APOL1 variant extraction*

Eight positions were extracted with bcftools[S3]: rs2239785 (chr22:36265284; p.E150K), rs136175 (chr22:36265520; p.M228I), rs136176 (chr22:36265600; p.R255K), rs73885316 (chr22:36265628; p.N264K, M1), rs73885319 (chr22:36265860; p.S342G, a G1 component), rs60910145 (chr22:36265988; p.I384M, a G1 component), rs71785313 (chr22:36265995; the G2 six base pair deletion, merged with rs143830837 in dbSNP), and rs12106505 (chr22:36259617; the former G2 proxy).

##### *G2 detection*

G2 is a 6-bp in-frame deletion that removes p.Asn388 and p.Tyr389. G2 was determined directly from this deletion at chr22:36265995, identified by its rsID or by a six base pair length difference between the reference and alternate alleles. For haplotype frequency distribution, G2 and its background haplotypes were directly called from phased sequences. The program used to estimate allele age requires biallelic input. For these analyses, the upstream variant rs12106505 was used as a G2 proxy. In ADSP R5, 197 participants (including 100 reported as Black or African American) carried the proxy allele without the G2 deletion, so proxy-based calling misclassifies a minority of participants in the allele age analyses.

##### *Haplotype classification*

Each phased haplotype was assigned a four-letter code (amino acids at positions 150, 228, 255, and 264), which identified the background protein haplotype (positions 150, 228, 255) and the presence or absence of the M1 modifier (position 264), and a risk suffix (G0, G1, or G2) (Supplementary Figure S1, Supplementary Table S1). G1 was defined as the alternate allele at both rs73885319 and rs60910145 on the same phased haplotype. G2 was the 6 base pair deletion on that haplotype. Because the data were phased, both haplotypes of each participant were read directly.

***Reported race and ethnicity variables in ADSP R5***

Reported race and reported ethnicity are separate ADSP phenotype variables. Per the ADSP data dictionary, both are to be described as reported rather than self-reported, because these variables were administratively assigned in some cohorts. Race is coded 1 = American Indian or Alaska Native, 2 = Asian, 3 = Native Hawaiian or Other Pacific Islander, 4 = Black or African American, 5 = White, and 6 = Other; ethnicity is coded 1 = Hispanic or Latino and 0 = Not Hispanic or Latino. A value was treated as usable only when the participant had a record in the ADSP R5 case-control phenotype file and the corresponding race or ethnicity field was not missing.

Of the 40,909 participants analyzed, 33,058 had a usable reported race and 31,719 a usable reported ethnicity. The 7,851 participants without a usable reported race comprise 7,761 whose subject identifier had no record in the case-control phenotype file and 90 who had a record but a missing race value; these participants were not assigned to any reported-race category and are reported throughout as a separate Unknown or no record stratum. The 9,190 participants without a usable reported ethnicity comprised the same 7,761 with no record in that file and 1,429 who had a record but a missing ethnicity value. The two sets are not nested: 1,405 participants had a usable reported race but no usable reported ethnicity and 66 the reverse, which is why the reported-ethnicity denominator in Supplementary Table S4 is 31,719 rather than 33,058. Ethnicity is recorded independently of race, and the 9,693 participants with reported Hispanic or Latino ethnicity fall across all six reported-race categories and the Unknown or no record stratum.

***Genetic ancestry and principal component analysis***

To evaluate the reported-race labels used in the main analysis, genetic ancestry was estimated independently with PLINK 2.0.[S4] Autosomal biallelic single-nucleotide variants were retained after variant-missingness (--geno 0.02) and Hardy-Weinberg ($p$ > 1E-6) filters; long-range linkage and major histocompatibility complex regions were excluded; and variants were pruned for pairwise correlation (--indep-pairwise 500 50 0.2). Unrelated participants were identified with KING[S5] (second-degree cutoff 0.0884).

Principal components were computed on the unrelated, pruned set (approximate algorithm, 10 components with allele weights), and all remaining participants were projected onto these axes.[S6,S7] Ten components were retained throughout, and different parts of the analysis use different numbers of them: the random-forest classifier and the African-ancestry-fraction estimation use PC1 to PC6, the model-based clustering uses PC1 to PC4, and the figures display PC1 and PC2. A 500-tree random forest[S8] trained on 1000 Genomes[S9] superpopulation labels assigned continental ancestry from PC1 to PC6. Assignments with a maximum class probability of at least 0.80 were considered confident; participants below this threshold were classified as Admixed/Unassigned. Participants with confident AFR assignments formed the AFR stratum (n = 4,455) and participants with confident AMR assignments formed the Admixed American stratum and served as a proxy for HIS in the figures (n = 8,998); participants assigned to EUR,

EAS, or SAS, and those classified as Admixed/Unassigned, were excluded from the AFR-and-HIS plots. Separately, constrained non-negative least squares over PC1 to PC6 and the five 1000 Genomes superpopulation centroids was used to estimate the African-ancestry fraction.

***Model-based clustering of genetic ancestry***

Model-based clustering of the principal components used the *mclust*[S10] package with the VVV model (varying volume, shape, and orientation, that is a full unconstrained covariance per cluster), applied to principal components 1 through 4. Four axes were used because four discriminant dimensions separate five reference superpopulations, so PC1 to PC4 is the smallest set that can resolve the 1000 Genomes reference structure. Because the Bayesian information criterion continues to improve with added clusters at this sample size and does not identify a natural cluster count, the number of clusters was fixed rather than selected by the Bayesian information criterion. Solutions with G = 5, 6, and 7 were compared directly. G = 7 was the smallest solution that recovered all five 1000 Genomes superpopulations as distinct clusters and was used as the primary solution. Cluster purity is the proportion of the 1000 Genomes reference samples falling in a cluster that carry that cluster's superpopulation label. G = 5 merged the Admixed American, South Asian, and East Asian groups, and G = 6 left East Asians within the Admixed American cluster.

The 1000 Genomes labels did not enter the mixture model likelihood and did not determine cluster membership. They were used after fitting for two purposes: to select the number of clusters, by taking the smallest G that resolved all five superpopulations as distinct clusters, and to name each cluster: for every cluster, the mean of its member samples in PC1 to PC4 space was compared with the centroid of each of the five 1000 Genomes superpopulations in the same space, and the cluster took the label of the superpopulation at the smallest Euclidean distance.

##### *Agreement between reported race and genetic ancestry*

Concordance between reported race or ethnicity and genetically inferred ancestry was quantified with Cohen kappa[S11,S12] (one versus rest), following the framework of Hall *et al*.[S13] Because this study focuses on African-ancestry populations, the African comparison (genetically inferred African versus reported Black or African American) is the primary agreement statistic. The full multi-group cross-tabulation is given in Supplementary Table S3, and agreement, sensitivity, and positive predictive value for each comparison are given in Supplementary Table S4.

##### *Relate allele-age estimation*

Local genealogies across an approximately 10 Mb window centered on *APOL1* were inferred with Relate v1.2.4[S14] and allele ages were sampled from the inferred genealogies. G2 was represented by the proxy single-nucleotide variant rs12106505 because G2 is a six base pair deletion and was not retained in the biallelic single-nucleotide input required for Relate. Allele ages were obtained from the population-size-calibrated genealogy assuming a generation time of 28 years. Because inference used a local window and a neutral coalescent model at a locus with evidence of selection, absolute ages are approximate and may be inflated; we therefore interpret lineage placement and relative ordering rather than exact calendar dates (Supplementary Figures S4 and S5).

##### *Ascertainment sensitivity*

To assess whether Alzheimer disease ascertainment distorts *APOL1* haplotype structure, phased haplotypes were linked by subject identifier both to the ADSP R5 case-control phenotype file and to the ADSP phenotype harmonization consortium demographics and diagnosis file (release 2026-02-18). The harmonized file records the last available diagnosis on a five-level scale, so it separates mild cognitive impairment from unimpaired and demented participants and assigns a status to many participants who have none in the binary case-control field. Cognitive status was therefore taken from the harmonized variable where available and from the binary field otherwise, giving three analysis groups (no cognitive impairment, mild cognitive impairment and Alzheimer dementia) and classifying 3,978 of the 4,371 participants in the primary stratum, against 3,243 with the binary field alone. Participants whose last diagnosis was non-AD dementia or another cognitive diagnosis were excluded by design, as were those with no diagnosis information in either file. Carrier frequencies of G1, G2 and M1, the frequency of the high-risk genotype, sex and *APOE* ε4 status were compared across the three groups with the Pearson chi-square test, or the Fisher exact test when an expected cell count was below 5; age was compared with the Kruskal-Wallis test (Supplementary Table S5).

***Software used***

bcftools; Beagle 5.5 (27 February 2025 build); PLINK 2.0; Relate v1.2.4; mclust; KING, R 4.5.1; Python 3.10. Analysis code is available from the corresponding author on request.

#### 2. APOL1 cation-flux experiments

***Cell generation***

Tetracycline regulated expression (T-REx)-293 cells were purchased (Thermofisher). *APOL1* plasmids with the indicated kidney risk variants and haplotypes were generated using QuikChange II Site-Directed Mutagenesis Kit (Agilent), purified with EndoFree Maxi-prep (Qiagen) and transfected into respective T-REx cells using FuGENE 4K (ProMega). Cells were maintained in culture according to manufacturer recommendations with DMEM + Glutamax (Gibco) supplemented with 10% tetracycline-free FBS (Gibco), 1% Pen-Strep, 5 μg/ml blasticidin, and 400 μg/ml zeocin. APOL1 expression was induced with tetracycline in sterile water at the specified doses.

***Lysate collection, gel electrophoresis and western blotting***

Cells were seeded at 6.0 x $10^5$ cells/well in six well plates coated with 50 μg/ml rat collagen type I overnight. Cells were stimulated with tetracycline 100 ng/ml for six hours and then washed twice with ice cold PBS. Lysates were collected in RIPA Buffer (50 mM Tris-HCl pH 7.5, 0.1% SDS, 0.5% DOC, 1% Triton X-100), triturated and incubated on ice for 15 minutes. Lysates were cleared by centrifuging 16,000 x g at 4C. Lysates were quantified using a DC protein assay (Bio-Rad). SDS-PAGE and western blot transfer was performed as previously described.[20] Immunoblotting was performed by blocking in 5% non-fat milk with TBS-tween-20 0.2% for 60 minutes. APOL1 (Proteintech, Catalog: 66124-1-Ig, 1:40000 in 5% non-fat milk TBS-tween-20 0.2%). Coomassie staining performed on PVDF by incubating Coomassie Red Staining Solution (0.1% Coomassie Brilliant Blue R-250, 45% methanol, 10% glacial acetic acid) for three minutes and destaining (10% methanol, 10% glacial acetic acid) until lanes visible. PVDF images were captured as 600x600 dpi TIFF using an iBright CL/FL 1500 (Thermofisher, Firmware version 1.8.1) using smart exposure settings.

***Tl+ flux assays***

The FluxOR II Green Potassium Ion Channel Assay (Thermofisher) was performed according to manufacturer's instructions.[S15,S16] Briefly, cells were plated on 96 well Costar black wall clear bottom plates, attached overnight and induced with tetracycline 100 ng/ml for 6 hours. Cells were incubated with loading buffer (Component C, Component D, FluxOR II Reagent, deionized water and 2.5 mM probenecid) for one hour covered at room temperature. Loading buffer was replaced with assay buffer (DI water, Component C, 1x Component I and 2.5 mM probenecid) containing inaxaplin 3 μM (when indicated). Background readings were recorded on a Synergy H1 microplate reader (BioTek) using Gen5 software (version 3.17, Agilent) software at 475/530 nm (ex/em) obtained every two seconds for 30 seconds, followed by addition of stimulus buffer

(2 mM thallium sulfate, Component F and DI water, with nigericin 20 μM, when indicated) and recording 475/530 nm (ex/em) reading every two seconds for 180 seconds. Individual experiments consisted of four wells per treatment, except nigericin stimulated control wells (2 wells). Data presented is background subtracted. Slopes were calculated using linear regression from fluorescence data measured during the initial 30 seconds after thallium addition. Normalized values of the slopes were calculated by dividing the rate of the stimulated condition by the rate of the unstimulated condition for each genotype-specific cell line. Two-way ANOVA with Tukey's Honestly significant Difference (HSD shown). All statistics performed in R (v4.5.1).[S17]

### Supplementary Tables

#### Supplementary Table S1. *APOL1* Variant Reference and Haplotype Nomenclature.

##### *(a) Coding Variants*

| Variant | rsID | Ensembl Annotation | Change | Role |
|---|---|---|---|---|
| p.E150K | rs2239785 | ENST00000397278.8:c.448G>A; ENSP00000380448.4:p.Glu150Lys | Glu to Lys | Defines KIK background (with p.M228I, p.R255K) |
| p.M228I | rs136175 | ENST00000397278.8:c.684G>A; ENSP00000380448.4:p.Met228Ile | Met to Ile | Background residue |
| p.R255K | rs136176 | ENST00000397278.8:c.764G>A; ENSP00000380448.4:p.Arg255Lys | Arg to Lys | Background residue |
| p.N264K (M1) | rs73885316 | ENST00000397278.8:c.792C>A; ENSP00000380448.4:p.Asn264Lys | Asn to Lys | M1 modifier |
| p.S342G (G1) | rs73885319 | ENST00000397278.8:c.1024A>G; ENSP00000380448.4:p.Ser342Gly | Ser to Gly | G1 risk allele (component) |
| p.I384M (G1) | rs60910145 | ENST00000397278.8:c.1152T>G; ENSP00000380448.4:p.Ile384Met | Ile to Met | G1 risk allele (component) |
| G2 | rs71785313 | ENST00000397278.8:c.1164_1169del; ENSP00000380448.4:p.Asn388_Tyr389del | 6-bp deletion (removes Asn388, Tyr389) | G2 risk allele |
| (G2 proxy) | rs12106505 | ENST00000397278.8:c.188-1979A>T; NC_000022.11:g.36259617A>T; no protein change | Upstream A/T | G2 proxy; for quality control |

##### *(b) Haplotype Backgrounds*

| Background | Residues at 150 / 228 / 255 | Description |
|---|---|---|
| EMR | Glu / Met / Arg | Reference haplotype, second most common outside Africa |
| EIK | Glu / Ile / Lys | Background on which the G1 and G2 risk alleles arose |
| KIK | Lys / Ile / Lys | Most common background; can carry the M1 modifier |

The four-letter haplotype code gives the amino acids at positions 150, 228, 255, and 264 of APOL1 for the Reference Sequence noted below; the fourth residue is Asn (N) on the ancestral chromosome and Lys (K) on the M1 (p.N264K) chromosome. A risk suffix (G0, G1, or G2) indicates the absence (G0) or presence (G1 or G2) of kidney risk variants. Ensembl identifiers (Ensembl release 116, GRCh38). Gene *APOL1*, ENSG00000100342; transcript ENST00000397278.8 (*APOL1*-202), which is both the Ensembl canonical transcript and the MANE Select transcript and corresponds to RefSeq NM_003661.4; protein ENSP00000380448.4 (398 amino acids). All coding (c.) and protein (p.) descriptions refer to transcript ENST00000397278.8, and all genomic positions are 1-based GRCh38 coordinates on the forward strand. Note that rs60910145 is triallelic: relative to the T reference allele, only the G allele encodes p.Ile384Met and contributes to G1, whereas the C allele is synonymous.

#### Supplementary Table S2. Distribution of *APOL1* Haplotypes in the ADSP R5 Genetically Inferred African Stratum.

| Haplotype | Genetically inferred African (N = 4,455; chromosomes = 8,910) n (%) | Reported Black or African American (N = 4,371; chromosomes = 8,742) n (%) |
|---|---|---|
| KIKN_G0 | 2 861 (32.11) | 2 973 (34.01) |
| EIKN_G0 | 2 240 (25.14) | 2 111 (24.15) |
| EIKN_G1 | 2 037 (22.86) | 1 935 (22.13) |
| EIKN_G2 | 1 102 (12.37) | 1 025 (11.73) |
| EMRN_G0 | 411 (4.61) | 458 (5.24) |
| KIKK_G0 | 136 (1.53) | 125 (1.43) |
| KIKK_G2 | 61 (0.68) | 59 (0.67) |
| EMKN_G0 | 60 (0.67) | 52 (0.59) |
| Other labeled haplotypes | 2 (0.02) | 4 (0.05) |

Each cell is the phased haplotype count (percentage of phased chromosomes in that column). The first column of counts is the genetically inferred African stratum, defined as the 4,455 participants that are genetically classified as AFR. The second column of counts is the reported Black or African American stratum. “Other labeled haplotypes” combines all remaining low-frequency labels.

#### Supplementary Table S3. Cross-Tabulation of Reported Race by Genetically Inferred Ancestry in ADSP R5.

| Reported race | AFR | AMR | EUR | EAS | SAS | Admixed/ Unassigned | Row total |
|---|---|---|---|---|---|---|---|
| American Indian or Alaska Native | 3 (2.3) | 66 (51.6) | 20 (15.6) | 1 (0.8) | 2 (1.6) | 36 (28.1) | 128 |
| Asian | 2 (0.1) | 48 (1.4) | 4 (0.1) | 697 (20.0) | 2 641 (75.7) | 98 (2.8) | 3 490 |
| Native Hawaiian or Pacific Islander | 0 (0.0) | 2 (28.6) | 2 (28.6) | 0 (0.0) | 0 (0.0) | 3 (42.9) | 7 |
| Black or African American | 3 911 (89.5) | 128 (2.9) | 24 (0.5) | 2 (0.0) | 1 (0.0) | 305 (7.0) | 4 371 |
| White | 51 (0.3) | 2 846 (14.9) | 14 120 (74.1) | 2 (0.0) | 3 (0.0) | 2 033 (10.7) | 19 055 |
| Other or more than one race | 226 (3.8) | 4 389 (73.1) | 58 (1.0) | 0 (0.0) | 5 (0.1) | 1 329 (22.1) | 6 007 |
| Unknown / no record | 262 (3.3) | 1 519 (19.3) | 5 154 (65.6) | 73 (0.9) | 26 (0.3) | 817 (10.4) | 7 851 |
| [Reported Hispanic or Latino ethnicity] | 347 (3.6) | 7 413 (76.5) | 231 (2.4) | 5 (0.1) | 0 (0.0) | 1 697 (17.5) | 9 693 |
| Column total | 4 455 | 8 998 | 19 382 | 775 | 2 678 | 4 621 | 40 909 |

Each cell gives the number of participants with that combination of reported race and genetically inferred ancestry, with the percentage of the reported-race row in parentheses; row counts sum to the row total and row percentages sum to 100% across the six genetic-ancestry columns apart from rounding. Reported race and ethnicity are ADSP phenotype variables and are described as reported rather than self-reported, since these variables were administratively assigned in some cohorts. Reported ethnicity is recorded independently of reported race, so the reported Hispanic or Latino ethnicity row (Ethnicity = 1) overlaps every race row above it; it is shown separately and is excluded from the column totals to avoid double counting. Unknown or no record denotes the 7,851 participants with no usable reported race, comprising 7,761 whose subject identifier had no record in the ADSP R5 case-control phenotype file and 90 who had a record but a missing race value; these participants were not allocated to any reported-race category and appear only in this row. The AFR column is the stratum used in Supplementary Table S2. AFR, African; AMR, Admixed American; EUR, European; EAS, East Asian; SAS, South Asian.

#### Supplementary Table S4. Agreement Between Reported Race or Ethnicity and Genetically Inferred Ancestry in ADSP R5.

| Stratum | | N | Kappa (SE) | 95% CI | Agreement (%) | Sensitivity (%) | PPV (%) |
|---|---|---|---|---|---|---|---|
| ***AFR: reported Black or African American versus genetic AFR*** | | | | | | | |
| Both sexes | | 33 058 | 0.900 (0.004) | (0.893, 0.908) | 97.8 | 93.3 | 89.5 |
| | Male | 12 801 | 0.895 (0.007) | (0.882, 0.908) | 98.1 | 92.3 | 88.9 |
| | Female | 20 257 | 0.902 (0.004) | (0.894, 0.911) | 97.6 | 93.7 | 89.7 |
| ***HIS: reported Hispanic or Latino versus genetic AMR*** | | | | | | | |
| Both sexes | | 31 719 | 0.813 (0.004) | (0.806, 0.820) | 92.6 | 99.0 | 76.5 |
| | Male | 12 236 | 0.815 (0.006) | (0.803, 0.827) | 93.4 | 98.7 | 75.7 |
| | Female | 19 483 | 0.811 (0.005) | (0.802, 0.819) | 92.1 | 99.1 | 76.9 |
| ***EUR: reported White versus genetic EUR*** | | | | | | | |
| Both sexes | | 33 058 | 0.701 (0.004) | (0.694, 0.709) | 84.7 | 99.2 | 74.1 |
| | Male | 12 801 | 0.707 (0.006) | (0.695, 0.719) | 85.2 | 99.1 | 76.4 |
| | Female | 20 257 | 0.696 (0.005) | (0.687, 0.705) | 84.5 | 99.4 | 72.5 |

Cohen kappa, computed one versus rest within each group, between reported race or ethnicity and genetically inferred ancestry, following the framework of Hall *et al.*[S13] Genetic ancestry is treated as the reference standard and the reported label as the test, so sensitivity = P(reported positive | genetic positive) and PPV = P(genetic positive | reported positive). Denominators differ between comparisons because participants without a usable label are excluded: the AFR and EUR comparisons use the 33,058 participants with a usable reported race (40,909 minus the 7,851 with no usable reported race), and the HIS comparison uses the 31,719 participants with a usable reported ethnicity (40,909 minus the 9,190 with no usable reported ethnicity). AFR, African; AMR, Admixed American; EUR, European; HIS, Hispanic or Latino; CI, confidence interval; PPV, positive predictive value; SE, standard error.

#### Supplementary Table S5. *APOL1* Risk Variants and Participant Characteristics by Cognitive Diagnosis in ADSP R5 Participants Reported as Black or African American.

| **Characteristic** | **No cognitive impairment (N = 2,689)** | **Mild cognitive impairment (N = 457)** | **Alzheimer dementia (N = 832)** | ***P*** | **Test** |
|---|---|---|---|---|---|
| Age, median [IQR] | 79 [72–84] | 76 [68–82] | 81 [75–87] | <0.001 | KW |
| Female, n (%) | 1 894 (70.4) | 304 (66.5) | 569 (68.4) | 0.173 | $\chi^2$ |
| *APOE* ε4 carrier, n/N (%) | 644/1 952 (33.0) | 225/456 (49.3) | 436/725 (60.1) | <0.001 | $\chi^2$ |
| G1 carrier, n (%) | 1 050 (39.0) | 162 (35.4) | 313 (37.6) | 0.306 | $\chi^2$ |
| G2 carrier, n (%) | 622 (23.1) | 126 (27.6) | 203 (24.4) | 0.112 | $\chi^2$ |
| M1 (p.N264K) carrier, n (%) | 114 (4.2) | 19 (4.2) | 35 (4.2) | 0.996 | $\chi^2$ |
| High risk genotype, n (%) | 361 (13.4) | 61 (13.3) | 97 (11.7) | 0.409 | $\chi^2$ |

Cognitive status is the participant's last available harmonized diagnosis in the ADSP phenotype harmonization consortium demographics and diagnosis file, which records no cognitive impairment, mild cognitive impairment, Alzheimer dementia, non-AD dementia, or other cognitive diagnosis. This classifies 3,978 of the 4,371 participants (91.0%) since some individuals do not have a confirmed diagnosis. Age is the age recorded at the participant's last visit with a diagnosis in the harmonized file where available and the age field of the case-control file otherwise. KW, Kruskal-Wallis test. Carrier denotes at least one copy of the allele, and the high-risk genotype is two *APOL1* risk alleles in any combination of G1 and G2; phased haplotypes are available for all 3,978 classified participants. Categorical variables were compared across the three groups with the Pearson chi-square test ($\chi^2$). IQR, interquartile range.

### Supplementary Figures S1-S5

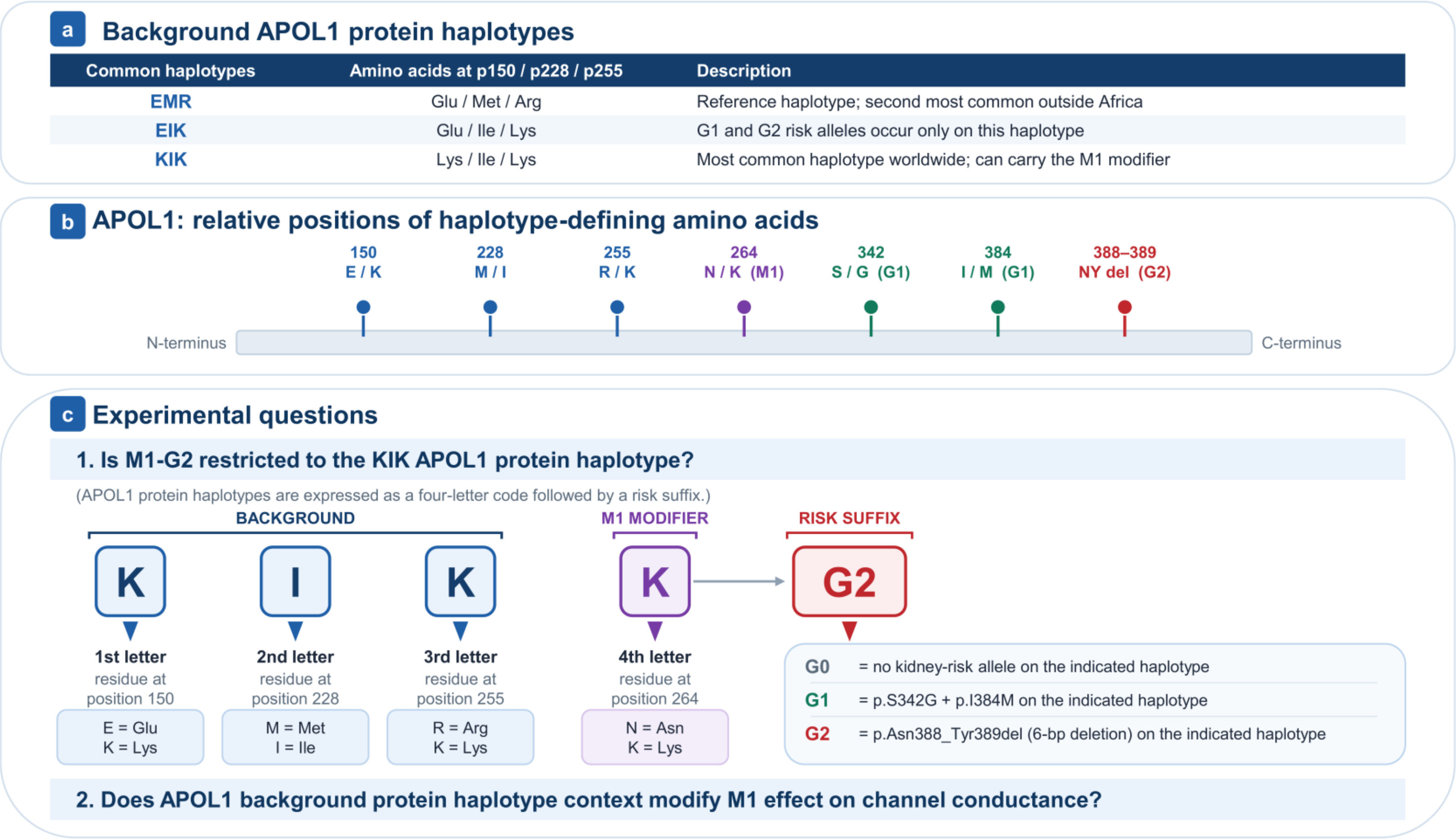

**Supplementary Figure S1. APOL1 protein haplotype-defining amino acids and experimental questions. a)** The background protein haplotypes discussed in this paper are defined by the amino acid sequence of APOL1 at three non-contiguous amino acid variations at positions 150, 228, and 255 of the reference sequence NM_003661.4, NP_003652.2. The single letter amino acid residues at these three positions of the reference sequence are EMR, which is the second most common APOL1 protein haplotype in non-African populations. The kidney disease associated risk variants G1 and G2 are carried on the EIK protein haplotype background. KIK is the most common haplotype worldwide. Recently a SNP encoding amino acid 264, the M1 modifier, was found to mitigate the kidney disease risk when co-inherited with the G2 kidney disease risk variant. To date, the M1 modifier has only been found on a non-risk G0 KIK haplotype or in *cis* with G2 on a KIK haplotype but has not been observed with the G1 kidney disease risk variant or other haplotypes. **b)** Relative positions of the APOL1 protein haplotype-defining amino acids, the M1 modifier and the kidney disease associated risk variants G1 and G2. GO lacks G1 or G2 risk variants. **c)** A template for the APOL1 protein haplotypes used in this paper is shown. *APOL1* variant reference and haplotype nomenclature is shown in Supplementary Table S1. To date, M1 containing APOL1 protein haplotypes have been identified in relatively small populations. This study has 2 goals. First, we characterized phased haplotype frequencies in the ADSP R5 (n=40,909 participants) and the non-Hispanic Black participants in All of Us (n=72,538) to determine if M1 is found on APOL1 protein haplotypes in addition to KIK. Second, we ask if the APOL1 haplotype modified the M1 block on G2 channel activity in HEK 293 cells stably expressing tetracycline-regulatable *APOL1* G2 transgenes with EIKN, EIKK, KIKN and KIKK haplotypes.

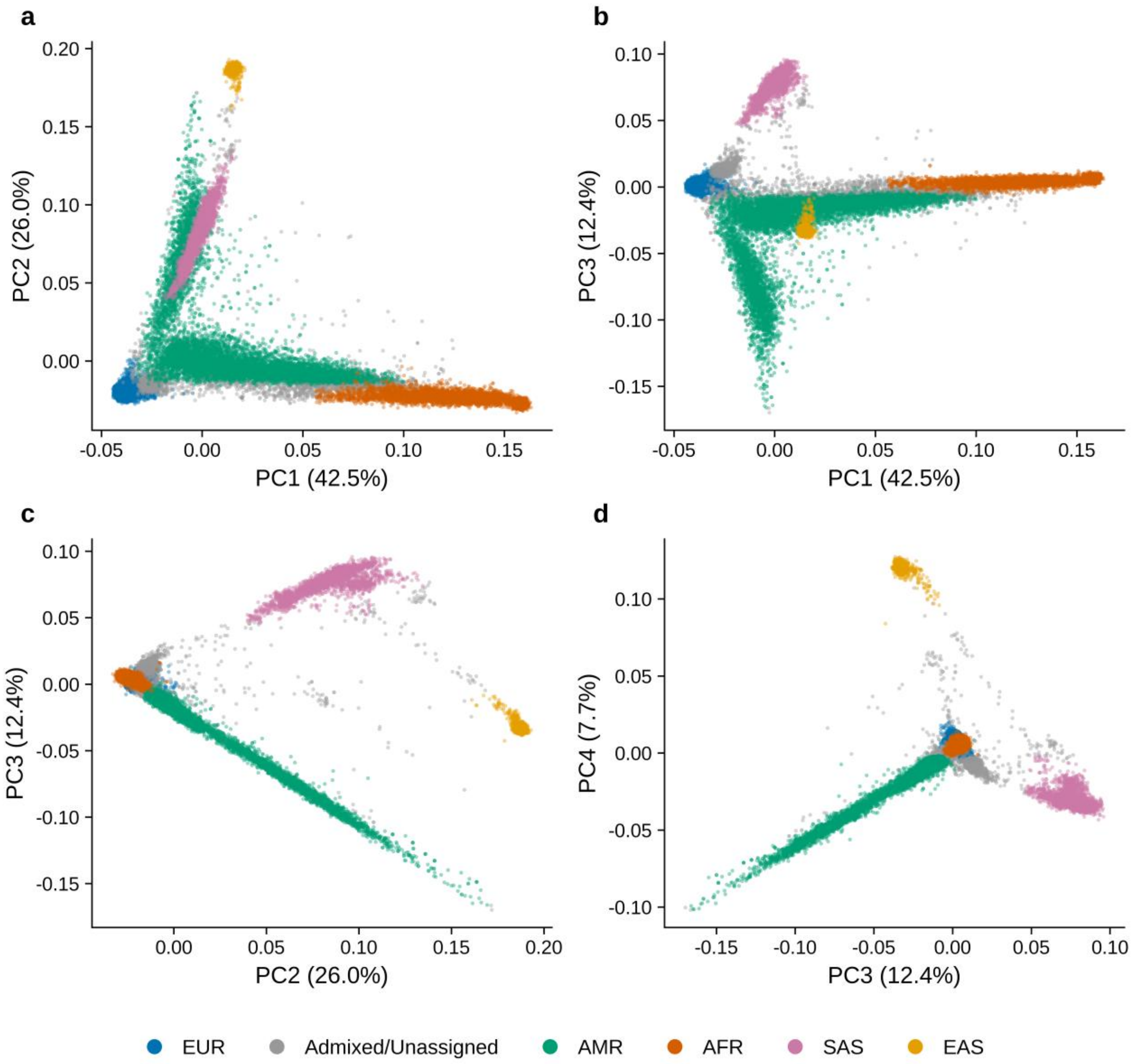

**Supplementary Figure S2. Unsupervised model-based clustering of ADSP participants.** Panels a to d show all participants projected onto pairs of the first four principal components (**a**, PC1 versus PC2; **b**, PC1 versus PC3; **c**, PC2 versus PC3; **d**, PC3 versus PC4). Model-based clustering was performed on principal components 1 to 4 using a seven-component VVV Gaussian mixture model; four axes were used because four discriminant dimensions separate the five 1000 Genomes reference superpopulations. The 1000 Genomes superpopulation labels were not used to fit the model and did not determine cluster membership; they were applied only after fitting, to name each cluster. Each cluster took the label of the 1000 Genomes superpopulation whose centroid in principal component 1 to 4 space lay at the smallest Euclidean distance from the cluster mean. AFR, African; AMR, Admixed American; EAS, East Asian; EUR, European; SAS, South Asian.

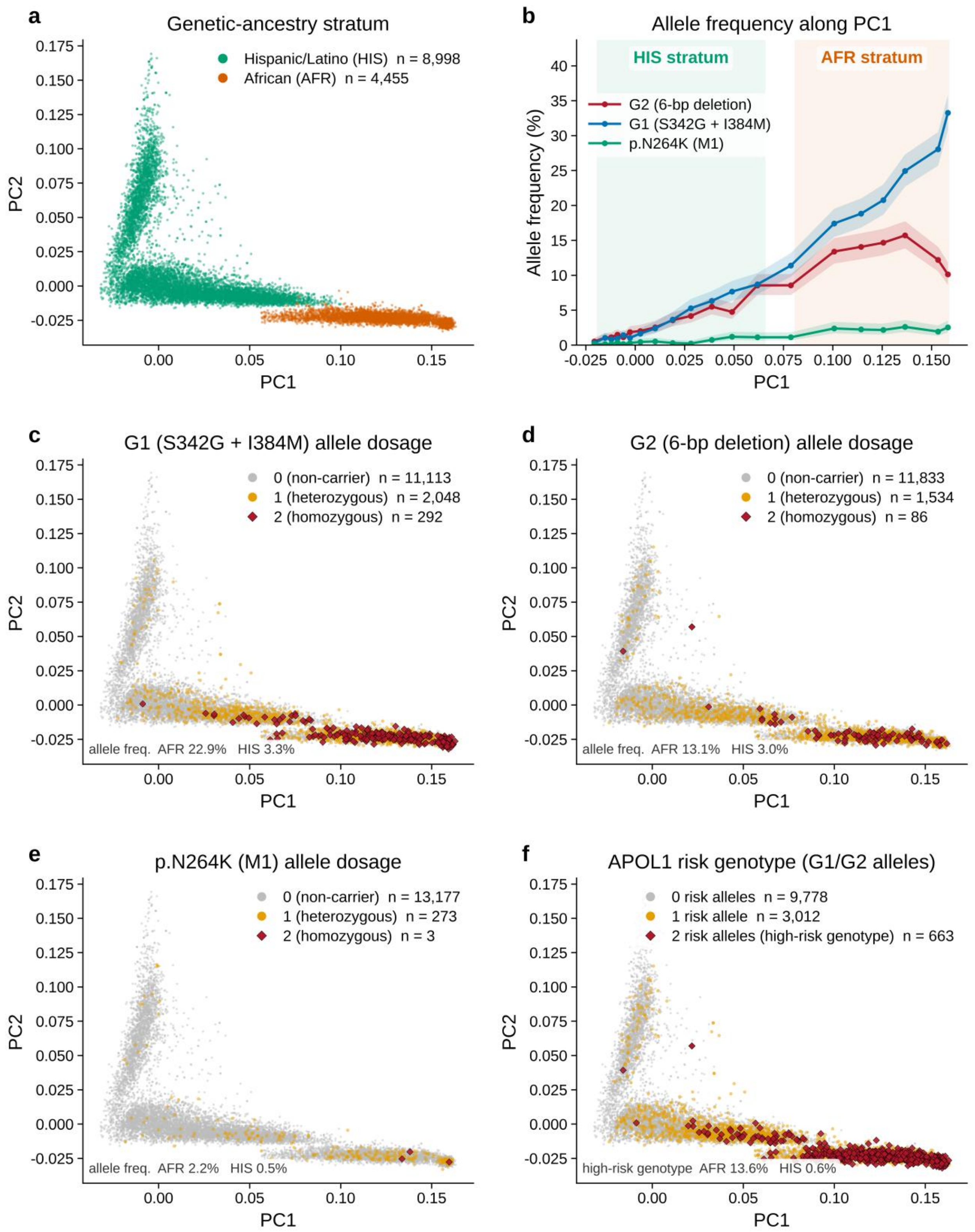

**Supplementary Figure S3. *APOL1* G1, G2 and p.N264K carriers in the genetic-ancestry principal-component space of the African and Hispanic or Latino strata.** Principal components 1 and 2 of the ADSP R5 ancestry principal-component analysis for the AFR (n =

4,455) and HIS (n = 8,998) genetic-ancestry strata, 13,453 participants in total. The Hispanic or Latino stratum is shown alongside the African stratum because participants in that stratum carry African ancestry in variable proportion, which extends the range of African-ancestry fraction along principal component 1 and is what makes the allele-frequency gradient in panel b measurable. (**a**) Genetic-ancestry stratum. (**b**) *APOL1* allele frequency along principal component 1. Participants were ranked by principal component 1 and divided into 20 equally sized groups (672 to 673 participants each); each point is plotted at the median principal component 1 value of its group and shaded bands are Wilson 95% confidence intervals. (**c to e**) Allele dosage for G1, G2 and the M1 modifier p.N264K: gray, non-carrier; amber, heterozygote; dark red diamond, homozygote. (f) *APOL1* risk genotype; two risk alleles in any combination (G1/G1, G1/G2 or G2/G2) constitute the high-risk genotype. G1 is p.S342G in *cis* with p.I384M, G2 is the six base pair deletion, and M1 is p.N264K.

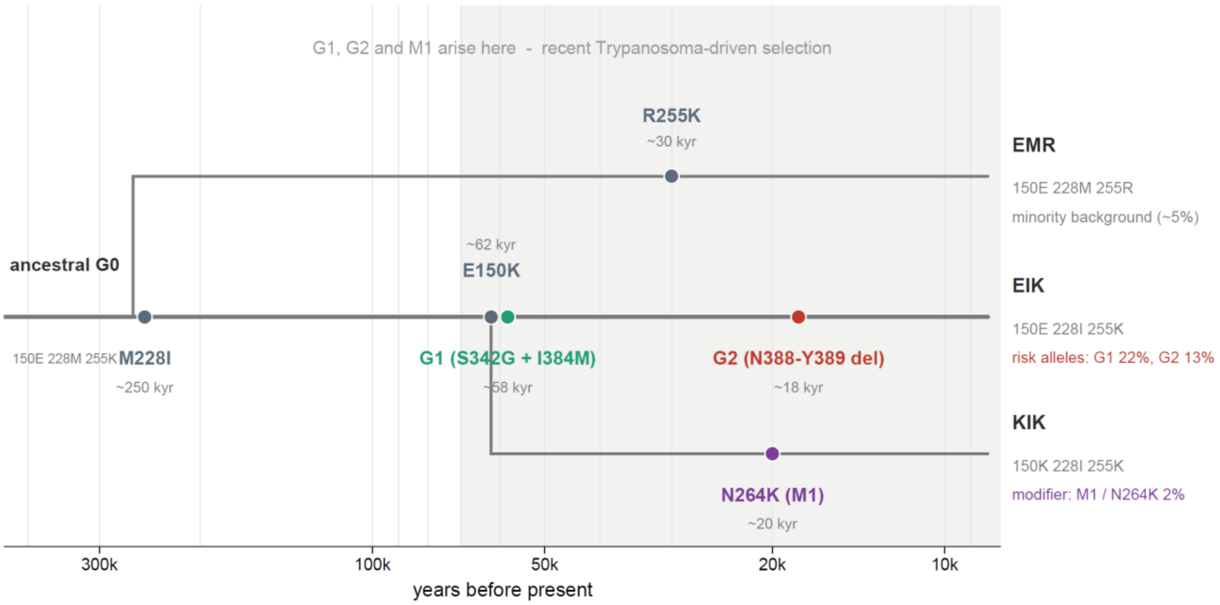

**Supplementary Figure S4. Estimated emergence of *APOL1* coding variants.** Schematic genealogy of the *APOL1* coding variants on the EMR, EIK, and KIK backgrounds, with approximate ages from the Relate analysis. The shaded region indicates the interval in which trypanosome-driven selection is proposed to have acted; the unshaded region indicates the earlier interval, which is not attributed to that selective pressure. p.M228I lies on the deepest focal branch, followed by p.E150K; p.N264K and the kidney risk alleles are recent relative to that older background substitution.

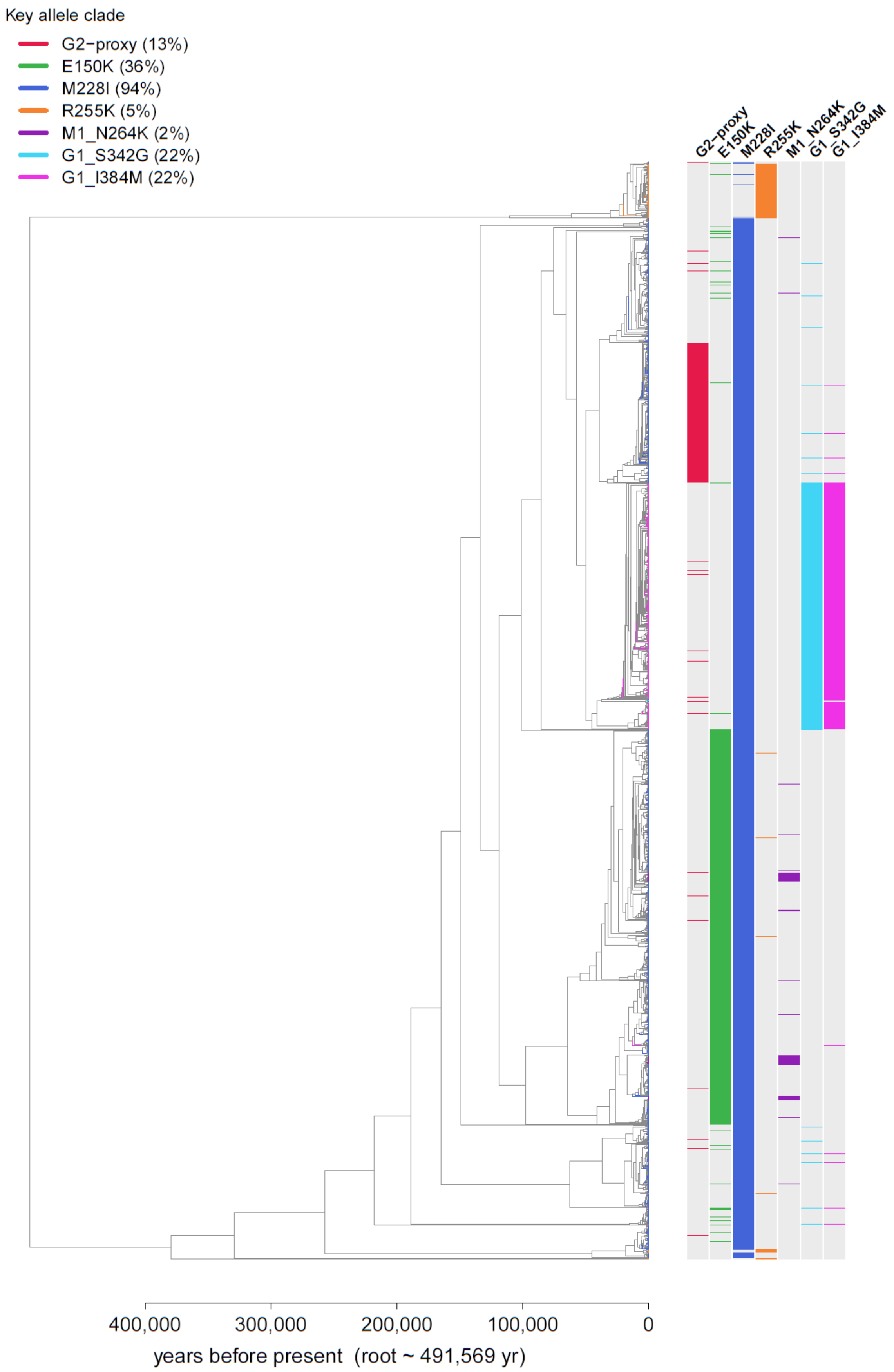
Key allele clade
G2−proxy (13%)
E150K (36%)
M228I (94%)
R255K (5%)
M1_N264K (2%)
G1_S342G (22%)
G1_I384M (22%)
G2−proxy
E150K
M228I
R255K
M1_N264K
G1_S342G
G1_I384M
400,000
300,000
200,000
100,000
0
years before present (root ~ 491,569 yr)

**Supplementary Figure S5. Relate-inferred local genealogy of *APOL1* haplotypes among ADSP R5 participants reported as Black or African American.** Local genealogy inferred with Relate across the *APOL1* region using phased haplotypes from ADSP R5 participants reported as Black or African American. Branch colors and the aligned tip tracks denote clades carrying the focal *APOL1* allele states; percentages in the key are their frequencies in this stratum, and the vertical axis is the Relate-inferred time scale in years before present. Chromosomes carrying p.N264K fall within the clade carrying p.E150K. G2 is represented by the upstream proxy variant rs12106505 because the six base pair deletion was not retained in the biallelic single-nucleotide input required for this analysis. Absolute ages are approximate; only lineage topology and relative ordering are interpreted.

#### Supplementary Acknowledgements

The Alzheimer's Disease Sequencing Project (ADSP) is comprised of two Alzheimer's Disease (AD) genetics consortia and three National Human Genome Research Institute (NHGRI) funded Large Scale Sequencing and Analysis Centers (LSAC). The two AD genetics consortia are the Alzheimer's Disease Genetics Consortium (ADGC) funded by NIA (U01 AG032984), and the Cohorts for Heart and Aging Research in Genomic Epidemiology (CHARGE) funded by NIA (R01 AG033193), the National Heart, Lung, and Blood Institute (NHLBI), other National Institute of Health (NIH) institutes and other foreign governmental and non-governmental organizations. The Discovery Phase analysis of sequence data is supported through UF1AG047133 (to Drs. Schellenberg, Farrer, Pericak-Vance, Mayeux, and Haines); U01AG049505 to Dr. Seshadri; U01AG049506 to Dr. Boerwinkle; U01AG049507 to Dr. Wijsman; and U01AG049508 to Dr. Goate and the Discovery Extension Phase analysis is supported through U01AG052411 to Dr. Goate, U01AG052410 to Dr. Pericak-Vance and U01 AG052409 to Drs. Seshadri and Fornage.

Sequencing for the Follow Up Study (FUS) is supported through U01AG057659 (to Drs. PericakVance, Mayeux, and Vardarajan) and U01AG062943 (to Drs. Pericak-Vance and Mayeux). Data generation and harmonization in the Follow-up Phase is supported by U54AG052427 (to Drs. Schellenberg and Wang). The FUS Phase analysis of sequence data is supported through U01AG058589 (to Drs. Destefano, Boerwinkle, De Jager, Fornage, Seshadri, and Wijsman), U01AG058654 (to Drs. Haines, Bush, Farrer, Martin, and Pericak-Vance), U01AG058635 (to Dr. Goate), RF1AG058066 (to Drs. Haines, Pericak-Vance, and Scott), RF1AG057519 (to Drs. Farrer and Jun), R01AG048927 (to Dr. Farrer), and RF1AG054074 (to Drs. Pericak-Vance and Beecham).

The ADGC cohorts include: Adult Changes in Thought (ACT) (U01 AG006781, U19 AG066567), the Alzheimer's Disease Research Centers (ADRC) (P30 AG062429, P30 AG066468, P30 AG062421, P30 AG066509, P30 AG066514, P30 AG066530, P30 AG066507, P30 AG066444, P30 AG066518, P30 AG066512, P30 AG066462, P30 AG072979, P30 AG072972, P30 AG072976, P30 AG072975, P30 AG072978, P30 AG072977, P30 AG066519, P30 AG062677, P30 AG079280, P30 AG062422, P30 AG066511, P30 AG072946, P30 AG062715, P30 AG072973, P30 AG066506, P30 AG066508, P30 AG066515, P30 AG072947,

P30 AG072931, P30 AG066546, P20 AG068024, P20 AG068053, P20 AG068077, P20 AG068082, P30 AG072958, P30 AG072959), the Chicago Health and Aging Project (CHAP) (R01 AG11101, RC4 AG039085, K23 AG030944), Indiana Memory and Aging Study (IMAS) (R01 AG019771), Indianapolis Ibadan (R01 AG009956, P30 AG010133), the Memory and Aging Project (MAP) ( R01 AG17917), Mayo Clinic (MAYO) (R01 AG032990, U01 AG046139, R01 NS080820, RF1 AG051504, P50 AG016574), Mayo Parkinson's Disease controls (NS039764, NS071674, 5RC2HG005605), University of Miami (R01 AG027944, R01 AG028786, R01 AG019085, IIRG09133827, A2011048), the Multi-Institutional Research in Alzheimer's Genetic Epidemiology Study (MIRAGE) (R01 AG09029, R01 AG025259), the National Centralized Repository for Alzheimer's Disease and Related Dementias (NCRAD) (U24 AG021886), the National Institute on Aging Late Onset Alzheimer's Disease Family Study (NIA- LOAD) (U24 AG056270), the Religious Orders Study (ROS) (P30 AG10161, R01 AG15819), the Texas Alzheimer's Research and Care Consortium (TARCC) (funded by the Darrell K Royal Texas Alzheimer's Initiative), Vanderbilt University/Case Western Reserve University (VAN/CWRU) (R01 AG019757, R01 AG021547, R01 AG027944, R01 AG028786, P01 NS026630, and Alzheimer's Association), the Washington Heights-Inwood Columbia Aging Project (WHICAP) (RF1 AG054023), the University of Washington Families (VA Research Merit Grant, NIA: P50AG005136, R01AG041797, NINDS: R01NS069719), the Columbia University Hispanic Estudio Familiar de Influencia Genetica de Alzheimer (EFIGA) (RF1 AG015473), the University of Toronto (UT) (funded by Wellcome Trust, Medical Research Council, Canadian Institutes of Health Research), and Genetic Differences (GD) (R01 AG007584). The CHARGE cohorts are supported in part by National Heart, Lung, and Blood Institute (NHLBI) infrastructure grant HL105756 (Psaty), RC2HL102419 (Boerwinkle) and the neurology working group is supported by the National Institute on Aging (NIA) R01 grant AG033193.

The CHARGE cohorts participating in the ADSP include the following: Austrian Stroke Prevention Study (ASPS), ASPS-Family study, and the Prospective Dementia Registry-Austria (ASPS/PRODEM-Aus), the Atherosclerosis Risk in Communities (ARIC) Study, the Cardiovascular Health Study (CHS), the Erasmus Rucphen Family Study (ERF), the Framingham Heart Study (FHS), and the Rotterdam Study (RS). ASPS is funded by the Austrian Science Fond (FWF) grant number P20545-P05 and P13180 and the Medical University of Graz.

The ASPS-Fam is funded by the Austrian Science Fund (FWF) project I904), the EU Joint Programme – Neurodegenerative Disease Research (JPND) in frame of the BRIDGET project (Austria, Ministry of Science) and the Medical University of Graz and the Steiermärkische Krankenanstalten Gesellschaft. PRODEM-Austria is supported by the Austrian Research Promotion agency (FFG) (Project No. 827462) and by the Austrian National Bank (Anniversary Fund, project 15435. ARIC research is carried out as a collaborative study supported by NHLBI contracts (HHSN268201100005C, HHSN268201100006C, HHSN268201100007C, HHSN268201100008C, HHSN268201100009C, HHSN268201100010C, HHSN268201100011C, and HHSN268201100012C). Neurocognitive data in ARIC is collected by U01 2U01HL096812, 2U01HL096814, 2U01HL096899, 2U01HL096902, 2U01HL096917 from the NIH (NHLBI, NINDS, NIA and NIDCD), and with previous brain MRI examinations funded by R01-HL70825 from the NHLBI. CHS research was supported by contracts HHSN268201200036C, HHSN268200800007C, N01HC55222, N01HC85079, N01HC85080, N01HC85081, N01HC85082, N01HC85083, N01HC85086, and grants U01HL080295 and U01HL130114 from the NHLBI with additional contribution from the National Institute of Neurological Disorders and Stroke (NINDS). Additional support was provided by R01AG023629, R01AG15928, and R01AG20098 from the NIA. FHS research is supported by NHLBI contracts N01-HC-25195 and HHSN268201500001I. This study was also supported by additional grants from the NIA (R01s AG054076, AG049607 and AG033040 and NINDS (R01 NS017950). The ERF study as a part of EUROSPAN (European Special Populations Research Network) was supported by European Commission FP6 STRP grant number 018947 (LSHG-CT-2006-01947) and also received funding from the European Community's Seventh Framework Programme (FP7/2007-2013)/grant agreement HEALTH-F4- 2007-201413 by the European Commission under the programme "Quality of Life and Management of the Living Resources" of 5th Framework Programme (no. QLG2-CT-2002- 01254). High-throughput analysis of the ERF data was supported by a joint grant from the Netherlands Organization for Scientific Research and the Russian Foundation for Basic Research (NWO-RFBR 047.017.043). The Rotterdam Study is funded by Erasmus Medical Center and Erasmus University, Rotterdam, the Netherlands Organization for Health Research and Development (ZonMw), the Research Institute for Diseases in the Elderly (RIDE), the Ministry of Education, Culture and Science, the Ministry for Health, Welfare and Sports, the European Commission (DG XII), and the municipality of Rotterdam. Genetic data sets are also supported by the Netherlands Organization of Scientific Research NWO Investments (175.010.2005.011, 911-03-012), the Genetic Laboratory of the Department of Internal Medicine, Erasmus MC, the Research Institute for Diseases in the Elderly (014-93-015; RIDE2), and the Netherlands Genomics Initiative (NGI)/Netherlands Organization for Scientific Research (NWO) Netherlands Consortium for Healthy Aging (NCHA), project 050-060-810. All studies are grateful to their participants, faculty and staff. The content of these manuscripts is solely the responsibility of the authors and does not necessarily represent the official views of the National Institutes of Health or the U.S. Department of Health and Human Services.

The FUS cohorts include: the Alzheimer's Disease Research Centers (ADRC) (P30 AG062429, P30 AG066468, P30 AG062421, P30 AG066509, P30 AG066514, P30 AG066530, P30 AG066507, P30 AG066444, P30 AG066518, P30 AG066512, P30 AG066462, P30 AG072979, P30 AG072972, P30 AG072976, P30 AG072975, P30 AG072978, P30 AG072977, P30 AG066519, P30 AG062677, P30 AG079280, P30 AG062422, P30 AG066511, P30 AG072946, P30 AG062715, P30 AG072973, P30 AG066506, P30 AG066508, P30 AG066515, P30 AG072947, P30 AG072931, P30 AG066546, P20 AG068024, P20 AG068053, P20 AG068077, P20 AG068082, P30 AG072958, P30 AG072959), Alzheimer's Disease Neuroimaging Initiative (ADNI) (U19AG024904), Amish Protective Variant Study (RF1AG058066), Cache County Study (R01AG11380, R01AG031272, R01AG21136, RF1AG054052), Case Western Reserve University Brain Bank (CWRUBB) (P50AG008012), Case Western Reserve University Rapid Decline (CWRURD) (RF1AG058267, NU38CK000480), CubanAmerican Alzheimer's Disease Initiative (CuAADI) (3U01AG052410), Estudio Familiar de Influencia Genetica en Alzheimer (EFIGA) (5R37AG015473, RF1AG015473, R56AG051876), Genetic and Environmental Risk Factors for Alzheimer Disease Among African Americans Study (GenerAAtions) (2R01AG09029, R01AG025259, 2R01AG048927), Gwangju Alzheimer and Related Dementias Study (GARD) (U01AG062602), Hillblom Aging Network (2014-A-004-NET, R01AG032289, R01AG048234), Hussman Institute for Human Genomics Brain Bank (HIHGBB) (R01AG027944, Alzheimer's Association "Identification of Rare Variants in Alzheimer Disease"), Ibadan Study of Aging (IBADAN) (5R01AG009956), Longevity Genes Project (LGP) and LonGenity (R01AG042188, R01AG044829, R01AG046949, R01AG057909, R01AG061155, P30AG038072), Mexican Health and Aging Study (MHAS) (R01AG018016),

Multi-Institutional Research in Alzheimer's Genetic Epidemiology (MIRAGE) (2R01AG09029, R01AG025259, 2R01AG048927), Northern Manhattan Study (NOMAS) (R01NS29993), Peru Alzheimer's Disease Initiative (PeADI) (RF1AG054074), Puerto Rican 1066 (PR1066) (Wellcome Trust (GR066133/GR080002), European Research Council (340755)), Puerto Rican Alzheimer Disease Initiative (PRADI) (RF1AG054074), Reasons for Geographic and Racial Differences in Stroke (REGARDS) (U01NS041588), Research in African American Alzheimer Disease Initiative (REAAADI) (U01AG052410), the Religious Orders Study (ROS) (P30 AG10161, P30 AG72975, R01 AG15819, R01 AG42210), the RUSH Memory and Aging Project (MAP) (R01 AG017917, R01 AG42210Stanford Extreme Phenotypes in AD (R01AG060747), University of Miami Brain Endowment Bank (MBB), University of Miami/Case Western/North Carolina A&T African American (UM/CASE/NCAT) (U01AG052410, R01AG028786), Wisconsin Registry for Alzheimer's Prevention (WRAP) (R01AG027161 and R01AG054047), Mexico-Southern California Autosomal Dominant Alzheimer's Disease Consortium (R01AG069013), Center for Cognitive Neuroscience and Aging (R01AG047649), and the A4 Study (R01AG063689, U19AG010483 and U24AG057437).

The four LSACs are: the Human Genome Sequencing Center at the Baylor College of Medicine (U54 HG003273), the Broad Institute Genome Center (U54HG003067), The American Genome Center at the Uniformed Services University of the Health Sciences (U01AG057659), and the Washington University Genome Institute (U54HG003079). Genotyping and sequencing for the ADSP FUS is also conducted at John P. Hussman Institute for Human Genomics (HIHG) Center for Genome Technology (CGT).

Biological samples and associated phenotypic data used in primary data analyses were stored at Study Investigators institutions, and at the National Centralized Repository for Alzheimer's Disease and Related Dementias (NCRAD, U24AG021886) at Indiana University funded by NIA. Associated Phenotypic Data used in primary and secondary data analyses were provided by Study Investigators, the NIA funded Alzheimer's Disease Centers (ADCs), and the National Alzheimer's Coordinating Center (NACC, U24AG072122) and the National Institute on Aging Genetics of Alzheimer's Disease Data Storage Site (NIAGADS, U24AG041689) at the University of Pennsylvania, funded by NIA. Harmonized phenotypes were provided by the

ADSP Phenotype Harmonization Consortium (ADSP-PHC), funded by NIA (U24 AG074855, U01 AG068057 and R01 AG059716) and Ultrascale Machine Learning to Empower Discovery in Alzheimer's Disease Biobanks (AI4AD, U01 AG068057). This research was supported in part by the Intramural Research Program of the National Institutes of Health, National Library of Medicine. Contributors to the Genetic Analysis Data included Study Investigators on projects that were individually funded by NIA, and other NIH institutes, and by private U.S. organizations, or foreign governmental or nongovernmental organizations.

The ADSP Phenotype Harmonization Consortium (ADSP-PHC) is funded by NIA (U24 AG074855, U01 AG068057 and R01 AG059716). The harmonized cohorts within the ADSP-PHC include: the Anti-Amyloid Treatment in Asymptomatic Alzheimer's study (A4 Study), a secondary prevention trial in preclinical Alzheimer's disease, aiming to slow cognitive decline associated with brain amyloid accumulation in clinically normal older individuals. The A4 Study is funded by a public-private-philanthropic partnership, including funding from the National Institutes of Health-National Institute on Aging, Eli Lilly and Company, Alzheimer's Association, Accelerating Medicines Partnership, GHR Foundation, an anonymous foundation and additional private donors, with in-kind support from Avid and Cogstate. The companion observational Longitudinal Evaluation of Amyloid Risk and Neurodegeneration (LEARN) Study is funded by the Alzheimer's Association and GHR Foundation. The A4 and A4-LEARN Studies are led by Dr. Reisa Sperling at Brigham and Women's Hospital, Harvard Medical School and Dr. Paul Aisen at the Alzheimer's Therapeutic Research Institute (ATRI), University of Southern California. The A4 and LEARN Studies are coordinated by ATRI at the University of Southern California, and the data are made available through the Laboratory for Neuro Imaging at the University of Southern California. The participants screening for the A4 Study provided permission to share their de-identified data in order to advance the quest to find a successful treatment for Alzheimer's disease. We would like to acknowledge the dedication of all the participants, the site personnel, and all of the partnership team members who continue to make the A4 and LEARN Studies possible. The complete A4 Study Team list is available on: a4study.org/a4-study-team.; the Adult Changes in Thought study (ACT), U01 AG006781, U19 AG066567; Alzheimer's Disease Neuroimaging Initiative (ADNI): Data collection and sharing for this project was funded by the Alzheimer's Disease Neuroimaging Initiative (ADNI) (National Institutes of Health Grant U01 AG024904) and DOD ADNI (Department of Defense award number W81XWH-12-2-0012). ADNI is funded by the National Institute on Aging, the National Institute of Biomedical Imaging and Bioengineering, and through generous contributions from the following: AbbVie, Alzheimer's Association; Alzheimer's Drug Discovery Foundation; Araclon Biotech; BioClinica, Inc.; Biogen; Bristol-Myers Squibb Company; CereSpir, Inc.; Cogstate; Eisai Inc.; Elan Pharmaceuticals, Inc.; Eli Lilly and Company; EuroImmun; F. Hoffmann-La Roche Ltd and its affiliated company Genentech, Inc.; Fujirebio; GE Healthcare; IXICO Ltd.;Janssen Alzheimer Immunotherapy Research & Development, LLC.; Johnson & Johnson Pharmaceutical Research & Development LLC.; Lumosity; Lundbeck; Merck & Co., Inc.; Meso Scale Diagnostics, LLC.; NeuroRx Research; Neurotrack Technologies; Novartis Pharmaceuticals Corporation; Pfizer Inc.; Piramal Imaging; Servier; Takeda Pharmaceutical Company; and Transition Therapeutics. The Canadian Institutes of Health Research is providing funds to support ADNI clinical sites in Canada. Private sector contributions are facilitated by the Foundation for the National Institutes of Health (www.fnih.org). The grantee organization is the Northern California Institute for Research and Education, and the study is coordinated by the Alzheimer's Therapeutic Research Institute at the University of Southern California. ADNI data are disseminated by the Laboratory for Neuro Imaging at the University of Southern California; Brain tissue and additional biological samples were collected from individuals who were assessed and followed in the Case Western ADC which was funded by NIA P50AG008012; Estudio Familiar de Influencia Genetica en Alzheimer (EFIGA): 5R37AG015473, RF1AG015473, R56AG051876; the Health & Aging Brain Study – Health Disparities (HABS-HD), supported by the National Institute on Aging of the National Institutes of Health under Award Numbers R01AG054073, R01AG058533, R01AG070862, P41EB015922, and U19AG078109; brain tissues were obtained from the Brain Bank at the John P. Hussman Institute for Human Genomics at the University of Miami Miller School of Medicine, with funding provided by Louis D. Scientific Prize from the Institut de France; the Indiana Alzheimer’s Disease Research Center (IADRC) supported by National Institutes of Health (NIH) grant P30AG072976; the Korean Brain Aging Study for the Early Diagnosis and Prediction of Alzheimer’s disease (KBASE), which was supported by a grant from Ministry of Science, ICT and Future Planning (Grant No: NRF-2014M3C7A1046042); Memory & Aging Project at Knight Alzheimer’s Disease Research Center (MAP at Knight ADRC): The Memory and Aging Project at the Knight-ADRC (Knight-ADRC). This work was supported by the

National Institutes of Health (NIH) grants R01AG064614, R01AG044546, RF1AG053303, RF1AG058501, U01AG058922 and R01AG064877 to Carlos Cruchaga. The recruitment and clinical characterization of research participants at Washington University was supported by NIH grants P30AG066444, P01AG03991, and P01AG026276. Data collection and sharing for this project was supported by NIH grants RF1AG054080, P30AG066462, R01AG064614 and U01AG052410. We thank the contributors who collected samples used in this study, as well as patients and their families, whose help and participation made this work possible. This work was supported by access to equipment made possible by the Hope Center for Neurological Disorders, the Neurogenomics and Informatics Center (NGI: https://neurogenomics.wustl.edu/) and the Departments of Neurology and Psychiatry at Washington University School of Medicine; the University of Miami Brain Endowment Bank (Miami Brain Bank); National Alzheimer's Coordinating Center (NACC): The NACC database is funded by NIA/NIH Grant U24 AG072122. SCAN is a multi-institutional project that was funded as a U24 grant (AG067418) by the National Institute on Aging in May 2020. Data collected by SCAN and shared by NACC are contributed by the NIA-funded ADRCs as follows: P30 AG062429 (PI James Brewer, MD, PhD), P30 AG066468 (PI Oscar Lopez, MD), P30 AG062421 (PI Bradley Hyman, MD, PhD), P30 AG066509 (PI Thomas Grabowski, MD), P30 AG066514 (PI Mary Sano, PhD), P30 AG066530 (PI Helena Chui, MD), P30 AG066507 (PI Marilyn Albert, PhD), P30 AG066444 (PI John Morris, MD), P30 AG066518 (PI Jeffrey Kaye, MD), P30 AG066512 (PI Thomas Wisniewski, MD), P30 AG066462 (PI Scott Small, MD), P30 AG072979 (PI David Wolk, MD), P30 AG072972 (PI Charles DeCarli, MD), P30 AG072976 (PI Andrew Saykin, PsyD), P30 AG072975 (PI David Bennett, MD), P30 AG072978 (PI Neil Kowall, MD), P30 AG072977 (PI Robert Vassar, PhD), P30 AG066519 (PI Frank LaFerla, PhD), P30 AG062677 (PI Ronald Petersen, MD, PhD), P30 AG079280 (PI Eric Reiman, MD), P30 AG062422 (PI Gil Rabinovici, MD), P30 AG066511 (PI Allan Levey, MD, PhD), P30 AG072946 (PI Linda Van Eldik, PhD), P30 AG062715 (PI Sanjay Asthana, MD, FRCP), P30 AG072973 (PI Russell Swerdlow, MD), P30 AG066506 (PI Todd Golde, MD, PhD), P30 AG066508 (PI Stephen Strittmatter, MD, PhD), P30 AG066515 (PI Victor Henderson, MD, MS), P30 AG072947 (PI Suzanne Craft, PhD), P30 AG072931 (PI Henry Paulson, MD, PhD), P30 AG066546 (PI Sudha Seshadri, MD), P20 AG068024 (PI Erik Roberson, MD, PhD), P20 AG068053 (PI Justin Miller, PhD), P20 AG068077 (PI Gary Rosenberg, MD), P20 AG068082 (PI Angela Jefferson, PhD), P30

AG072958 (PI Heather Whitson, MD), P30 AG072959 (PI James Leverenz, MD); National Institute on Aging Alzheimer's Disease Family Based Study (NIA-AD FBS): U24 AG056270; Religious Orders Study (ROS): P30AG10161,R01AG15819, R01AG42210; Memory and Aging Project (MAP - Rush): R01AG017917, R01AG42210; Minority Aging Research Study (MARS): R01AG22018, R01AG42210; the Texas Alzheimer's Research and Care Consortium (TARCC), funded by the Darrell K Royal Texas Alzheimer's Initiative, directed by the Texas Council on Alzheimer's Disease and Related Disorders; Washington Heights/Inwood Columbia Aging Project (WHICAP): RF1 AG054023;and Wisconsin Registry for Alzheimer's Prevention (WRAP): R01AG027161 and R01AG054047. Additional acknowledgments include the National Institute on Aging Genetics of Alzheimer's Disease Data Storage Site (NIAGADS, U24AG041689) at the University of Pennsylvania, funded by NIA.

S17. R Core Team. R: A Language and Environment for Statistical Computing. R Foundation for Statistical Computing; 2025.
